# Altered blood cell traits underlie a major genetic locus of severe COVID-19

**DOI:** 10.1101/2020.09.09.20191700

**Authors:** Jingqi Zhou, Yitang Sun, Weishan Huang, Kaixiong Ye

## Abstract

**Purpose:** The genetic locus 3p21.31 has been associated with severe coronavirus disease 2019 (COVID-19), but the underlying pathophysiological mechanism is unknown.

**Methods:** To identify intermediate traits of the COVID-19 risk variant, we performed a phenome-wide association study (PheWAS) with 923 phenotypes in 310,999 European individuals from UK Biobank. For candidate target genes, we examined associations between their expression and the polygenic score (PGS) of 1,263 complex traits in a meta-analysis of 31,684 blood samples.

**Results:** Our PheWAS identified and replicated multiple blood cell traits to be associated with the COVID-19 risk variant, including monocyte count and percentage (*p* = 1.07×10^−8^, 4.09×10^−13^), eosinophil count and percentage (*p* = 5.73×10^−3^, 2.20×10^−3^), and neutrophil percentage (*p* = 3.23×10^−3^). The PGS analysis revealed positive associations between the expression of candidate genes and genetically predicted counts of specific blood cells: *CCR3* with eosinophil and basophil (*p* = 5.73×10^−21^, 5.08×10^−19^); *CCR2* with monocytes (*p* = 2.40×10^−10^); and *CCR1* with monocytes and neutrophil (*p* = 1.78×10^−6^, 7.17×10^−5^).

**Conclusions:** Multiple blood cell traits, especially monocyte, eosinophil, and neutrophil numbers, are associated with the COVID-19 risk variant and the expression of its candidate target genes, representing probable mechanistic links between the genetic locus 3p21.31 and severe COVID-19.

## INTRODUCTION

The coronavirus disease 2019 (COVID-19), caused by infection of the severe acute respiratory syndrome coronavirus 2 (SARS-CoV-2), affects individuals differently, with clinical manifestations ranging from asymptomatic infection, to mild flu-like symptoms, to severe respiratory failure^1–3^. Besides demographic factors and pre-existing conditions, genetic variation is partly responsible for these varying individual responses^4–6^. The first genome-wide association study (GWAS) for COVID-19 in patients with respiratory failure identified genetic associations at locus 3p21.31^4^, which was independently replicated by the COVID-19 Host Genetics Initiative^5^. The peak signal at this locus spans multiple chemokine receptor genes (e.g., *CCR9, CXCR6, XCR1* and *CCR1*) and risk variants are associated with the expression of *CXCR6, CCR1* and *SLC6A20^4^*. However, the underlying causal variant, the target gene, and the pathophysiological process are unknown.

Phenome-wide association study (PheWAS) is an unbiased approach that evaluates the associations of a disease-associated genetic variant (e.g., a COVID-19 risk variant) with a wide range of phenotypes (i.e., the phenome). PheWAS may identify intermediate traits or biomarkers residing in the causal physiological route from the genetic variant to the disease of interest. It may also reveal unexpected comorbidities that indicate shared biological mechanisms^7,8^. Similarly, expression quantitative trait locus (eQTL) analysis for a trait-associated genetic variant across the transcriptome can identify candidate causal genes that are either close (in *cis*) or remote (in *trans*) to the variant^9,10^. From the perspective of a candidate gene, insights could be gained into its physiological pathways and downstream functional effects by examining associations of its expression level with phenotypes across the phenome, or even with the genetically predicted phenotypic status if measured ones are unavailable^10^.

This project aimed to explore the mechanistic link between the genetic locus 3p21.31 and severe COVID-19. We first leveraged the deep phenotyping information in the UK Biobank (N = 310,999) and performed a PheWAS for a COVID-19 risk variant with 923 disease phenotypes, biomarkers and blood cell traits. Candidate associations were further replicated in an additional dataset. Moreover, for genes potentially regulated by the COVID-19 risk variant, associations between their expression levels and the polygenic scores (PGS) of 1,263 traits were evaluated in 31,684 blood samples. These two unbiased phenome-wide approaches converged on blood cell traits, especially counts of monocyte, eosinophil and neutrophil, as the possible intermediate link between the genetic locus 3p21.31 and severe COVID-19.

## MATERIALS AND METHODS

### Ethics statement

UK Biobank is a large population-based prospective study that recruited more than 500,000 individuals aged 40-69 years between 2006 and 2010. It was approved by the North West Multi-Centre Research Ethics Committee and proper informed consent was obtained. All participants received baseline measurements, donated biological materials, and provided access to their medical records^11^. Data for this project was accessed through an approved application to UK Biobank (Application ID: 48818).

### Phenome-wide association study for the COVID-19 risk variant

#### Discovery analysis

This was performed with data from UK Biobank. Only participants fulfilling the following criteria were included: 1) genetic ancestry is Caucasian; 2) included in the genetic principal component analysis; 3) not outliers for heterogeneity and missing genotype rate; 4) no sex chromosome aneuploidy; 5) self-reported sex matching genetic sex; 6) no high degree of genetic kinship, and 7) for relative pairs (kinship coefficient > 0.0884), a minimum number of participants were removed so that all those remaining are unrelated. A total of 310,999 unrelated individuals passed this quality control and filtering procedure.

Three sets of phenotypes were examined in our PheWAS: binary disease outcomes, blood and urine biomarkers, and blood cell traits. Binary disease status was defined by mapping ICD9/ICD10 diagnosis codes in the hospital episode statistics to phecodes in the PheCODE grouping system^12^. A total of 858 phecodes with case number no less than 200 were retained in our analysis (Supplementary Table 1). For continuous traits, our PheWAS included 34 blood and urine biochemistry markers, and 31 blood cell traits (Supplementary Table 2)^11^. Statistical association analyses were performed with scripts in R language and the PheWAS package^13^. Logistic regression was performed for binary disease outcomes and linear regression for continuous blood and urine biomarkers, adjusting for age, sex, genotyping array, assessment center, and the first 10 genetic principal components. We applied Bonferroni correction for the total number of phenotypes tested, although we note that this is a conservative approach because the phenotypes are not independent. Summary statistics for nominally significant associations between the COVID-19 risk variant and blood cell traits were also retrieved from GeneATLAS, a large database of associations based on 452,264 UK Biobank White British individuals^14^.

### Replication analysis

For blood cell traits that are associated with the COVID-19 risk variant at the nominal significance level in the discovery analysis, we tested if they could be replicated in another existing study. It was a GWAS meta-analysis for 36 blood cell traits in 173,480 European ancestry individuals across three cohorts: UK Biobank (N = 87,265), UK BiLEVE (N = 45,694), and INTERVAL (N = 40,521)^15^. Summary statistics for this study were retrieved from the IEU OpenGWAS database^16^.

### eQTL and polygenic score association analysis

To identify genes whose expression levels are associated with the COVID-19 risk variant, we inquired eQTL analysis results from GTEx and eQTLGen^10,17^. The GTEx project studies tissue-specific gene expression and regulation in 54 non-diseased tissue sites from about 1,000 individuals^17^. The eQTLGen Consortium conducted eQTL meta-analysis in 31,684 samples of blood and peripheral blood mononuclear cells from 37 datasets^10^. It also performed PGS analysis to evaluate the associations between the expression level of most genes and the polygenic scores of 1,263 traits^10^. The majority of samples in both studies are of European ancestry. In the PGS analysis, multiple PGS were calculated for each trait with different GWAS, sample ancestry, and *p*-value cutoffs (*p* = 0.01, 1×10^−3^, 1×10^−4^, 1×10^−5^, 5×10^−8^). For blood cell traits, three previous GWAS were used and designated as study 1^18^, study 2^19^, and study 3^20^, respectively. Statistical significance was defined with the false discovery rate approach (FDR < 0.05)^10^.

### Resources

eQTLGen: https://www.eqtlgen.org/index.htmlGene ATLAS: http://geneatlas.roslin.ed.ac.uk/GTEx: https://www.gtexportal.org/home/The COVID-19 GWAS Results Browser: https://ikmb.shinyapps.io/COVID-19_GWAS_Browser/The COVID-19 Host Genetics Initiative: https://www.covid19hg.org/The IEU OpenGWAS database: https://gwas.mrcieu.ac.uk/

## RESULTS

### The severe COVID-19 risk variant is associated with blood cell traits

The severe COVID-19 risk variant examined in this study is rs67959919 (G/A), whose risk allele A has an odds ratio (OR) of 2.07 (95% confidence interval (CI): 1.66-2.56, *p* × 4.69×10^−11^) for severe COVID-19 after adjustment for genetic principal components, age and sex^4^. It is in perfect linkage disequilibrium (LD, r^2^ = 1) with the lead variant, rs11385942 (A/GA, OR = 2.11, 95% CI: 1.70-2.61, *p* = 9.46×10^−12^) in European populations^21^. The lead variant is an insertion-deletion polymorphism and is not found in some existing datasets. To identify phenotypes associated with rs67959919, we performed a PheWAS in a subset of 310,999 unrelated European individuals from the UK Biobank after quality control and filtering (Supplementary Table 3 for baseline characteristics). A total of 923 phenotypes were investigated, including 858 binary disease outcomes, 34 blood and urine biomarkers, and 31 blood cell traits (Fig. 1, Supplementary Tables 1 and 2).

**Figure 1.**
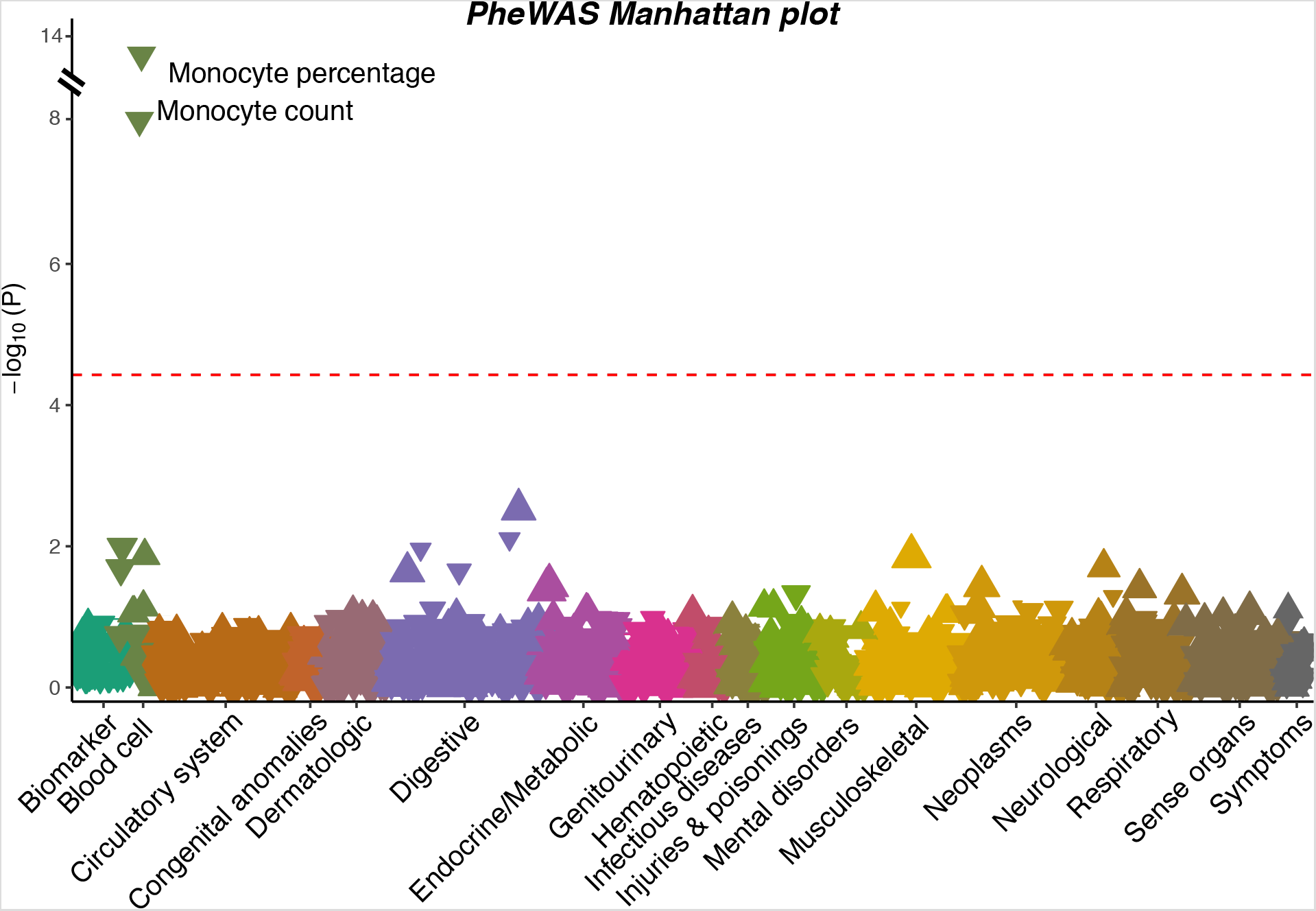
A Manhattan plot showing the associations between the severe COVID-19 risk variant and 923 phenotypes in UK Biobank. Each triangle represents one phenotype. Triangles pointing up indicate increasing effects of the COVID-19 risk allele on the phenotypes, while those pointing down indicate decreasing effects. The size of the triangle is proportional to the effect size. The significance threshold with Bonferroni correction (*p* < 0.05 / 923 = 5.42×10^−5^) is represented by the red dashed line.

With the conservative Bonferroni correction for the total number of phenotypes tested (*p* < 5.42×10^−5^), we observed that the severe COVID-19 risk variant is associated with monocyte percentage (*p* = 4.09×10^−13^) and monocyte count (*p* = 1.07×10^−8^). None of the binary disease outcomes or biomarkers passes this significance cutoff. The top three binary phenotypes were all related to the digestive system: sialolithiasis (*p* = 4.76×10^−4^), periodontitis (*p* = 1.53×10^−3^) and its subcategory, chronic periodontitis (*p* = 2.43×10^−3^). At the nominal significance level (*p* < 0.05), associations were observed with additional blood cell traits (Table 1): eosinophil count and percentage, neutrophil count and percentage, mean corpuscular hemoglobin, and mean corpuscular hemoglobin concentration. In GeneATLAS, a large database of associations based on 452,264 UK Biobank White British individuals^14^, consistent and even more significant associations were observed (Supplementary Table 4): monocyte percentage (*p* = 8.79×10^−26^), monocyte count (*p* = 1.15×10^−21^), eosinophil count (*p* = 3.81×10^−6^), eosinophil percentage (*p* = 1.54×10^−5^), neutrophil percentage (*p* = 1.03×10^−5^), basophil count (*p* = 1.40×10^−3^), basophil percentage (*p* = 1.87×10^−3^), red blood cell (RBC) count (*p* = 0.022), mean corpuscular hemoglobin (*p* = 0.023), and neutrophil count (*p* = 0.035). Notably, most of these associations were replicated in another study of 173,480 European-ancestry individuals (Table 1)^15^.

**Table 1:** Associations between the COVID-19 risk variant (rs67959919) and blood cell traits

| Trait | Discovery (UK Biobank)<br>N = 310,999 |  |  | Replication (Meta-analysis of three UK cohorts)<br>N = 173,480 |  |  |
| --- | --- | --- | --- | --- | --- | --- |
|  | beta | SE | *p* | beta | SE | *p* |
| Monocyte count | -0.0038 | 6.61e-4 | 1.07e-08 | -0.042 | 7.01e-3 | 2.34e-09 |
| Monocyte percentage | -0.0048 | 6.63e-4 | 4.09e-13 | -0.045 | 6.99e-3 | 1.45e-10 |
| Eosinophil count | -0.0018 | 6.52e-4 | 5.73e-03 | -0.024 | 7.00e-3 | 6.29e-04 |
| Eosinophil percentage | -0.0020 | 6.52e-4 | 2.20e-03 | -0.024 | 6.99e-3 | 6.66e-04 |
| Neutrophil count | 0.0014 | 6.50e-4 | 0.032 | 0.0042 | 7.04e-3 | 0.55 |
| Neutrophil percentage | 0.0019 | 6.50e-4 | 3.23e-03 | 0.015 | 7.01e-3 | 0.036 |
| Mean corpuscular hemoglobin | 0.0013 | 6.56e-4 | 0.042 | 0.014 | 6.95e-3 | 0.039 |
| Mean corpuscular hemoglobin concentration | 0.0013 | 6.61e-4 | 0.042 | -0.0055 | 6.80e-3 | 0.42 |

NOTE: The effect allele is the COVID-19 risk allele.

### Expression of candidate target genes is associated with PGS of blood cell traits

Candidate target genes of the COVID-19 risk variant through regulation of gene expression could be identified with eQTL analysis. Based on *cis*-eQTL analysis in 54 tissues from GTEx^17^, the COVID-19 risk variant is associated with the expression of *CXCR6, SLC6A20, CCR1, CCR9, RP11–697K23.3*, and *LZTFL1* in a total of 9 tissues (Supplementary Table 5). Moreover, eQTLGen^10^, a meta-analysis for *cis*-eQTL in 31,684 blood samples additionally identified the following genes: *FLT1P1, CCR3, SACM1L, CCR5, CCR2* and *RP11–24F11.2* (Supplementary Table 6). *Trans*-eQTL analysis in both studies did not identify any genes.

For all these candidate target genes of the COVID-19 risk variant, we interrogated if their expression levels are associated with the PGS of 1,263 traits examined in eQTLGen. A significant association indicates that the gene is implicated in pathways contributing to the trait^10^. Multiple significant associations were identified after correction for multiple testing (Fig. 2, Supplementary Table 7). Genetically predicted higher monocyte count is positively associated with the expression of *CCR1* (*p* = 1.78×10^−6^) and *CCR2* (*p* = 2.40×10^−10^). Genetically predicted higher eosinophil count and basophil count are positively associated with *CCR3* expression (*p* = 5.73×10^−21^ and *p* = 5.08×10^−19^, respectively). At the nominal significance level (*p* < 0.05), additional associations were observed for neutrophils, lymphocytes, RBC, and white blood cells (WBC). Predicted neutrophil count is positively associated with the expression of *CCR1* (*p* = 7.17×10^−5^), *FLT1P1* (*p* = 1.46×10^−5^), *SACM1L* (*p* = 5.69×10^−3^) and *CCR3* (*p* = 0.011). A negative association was observed between predicted lymphocyte count and *CCR1* (*p* = 9.41×10^−4^). Genetically predicted higher RBC count is associated with lower expression of *CXCR6* (*p* = 3.74×10^−4^) and *RP11-24F11.2* (*p* = 2.09×10^−3^), while for WBC count, a positive association was observed with *SACM1L* (*p* = 8.26×10^−4^). Notably, these associations were consistent across different GWAS datasets and *p*-value cutoffs used in the PGS calculation (Supplementary Table 7). These results suggest a possibility that the target gene of the COVID-19 risk variant is involved in hematologic processes and regulates blood cell counts.

**Figure 2.**
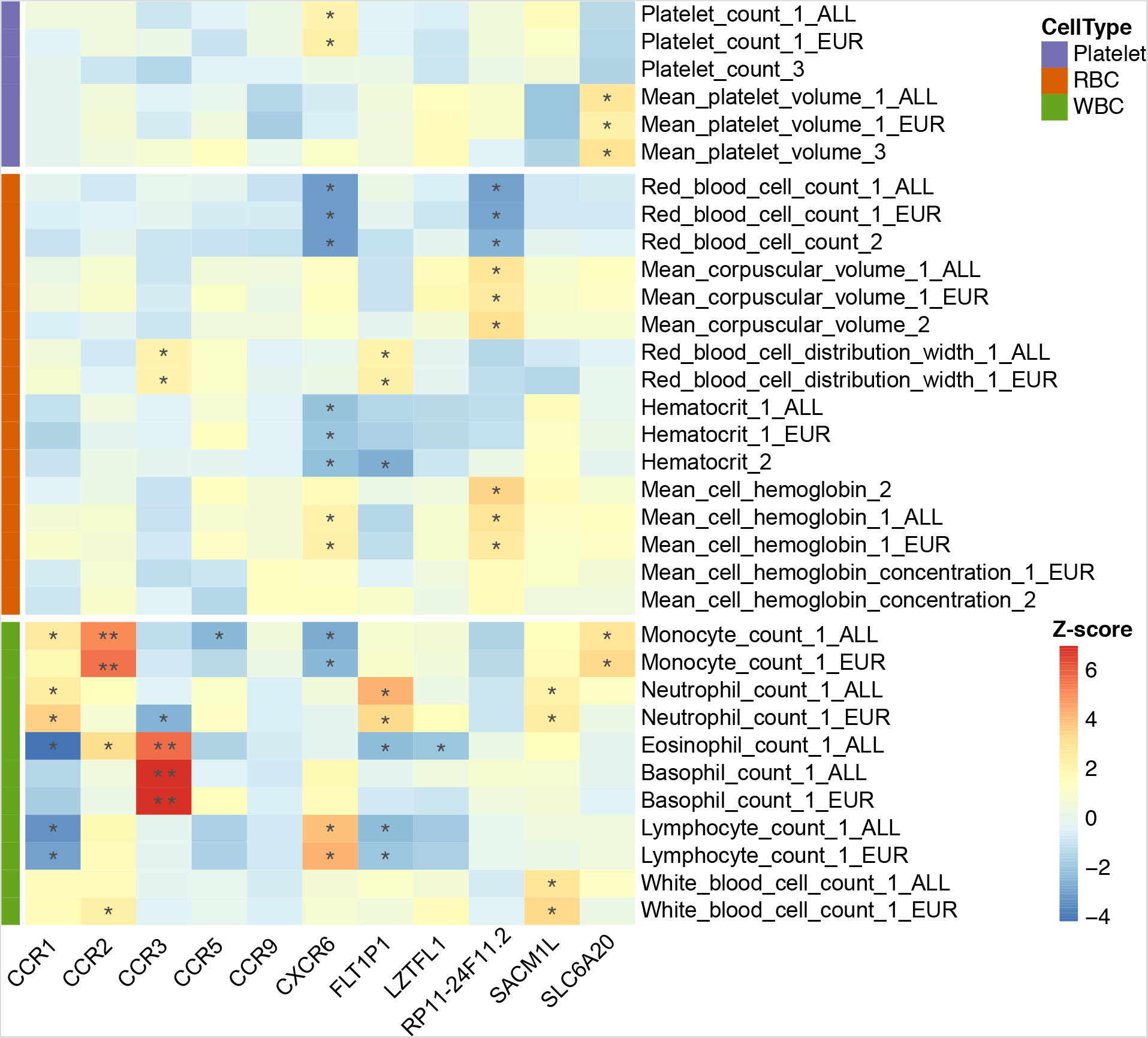
Associations between the expression of candidate genes and the polygenic score of blood cell traits. Each column corresponds to a gene. Each row corresponds to a polygenic score of a blood cell trait. Row names are organized as the combination of trait name, study number for the GWAS providing summary statistics, and the sample ancestry. EUR refers to European ancestry, while ALL refers to multi-ancestry. If no ancestry label is present, the study used only European samples. All PGS shown in this figure were calculated with a *p*-value cut off of 5×10^−8^. Complete association results for PGS calculated with other *p*-value cutoffs could be found in Supplementary Table 7. Blood cell traits are categorized into three groups: platelet, red blood cells, and white blood cells. The effects of association, Z-score, are shown as the heatmap. The statistical significance is indicated with “*” (p < 0.05) or “**” (FDR < 0.05).

Integrating and reconciling association signals in PheWAS, eQTL, and PGS analysis, three candidate blood cell traits and their corresponding candidate genes were prioritized (Fig. 3). First, the severe COVID-19 risk allele inhibits the expression of *CCR1* and *CCR2*, subsequently reducing the monocyte count. Second, the risk allele downregulates *CCR3* expression and further diminishes the eosinophil count. Third, the risk allele downregulates *CCR3* expression and relieves its inhibition on the neutrophil count.

**Figure 3.**
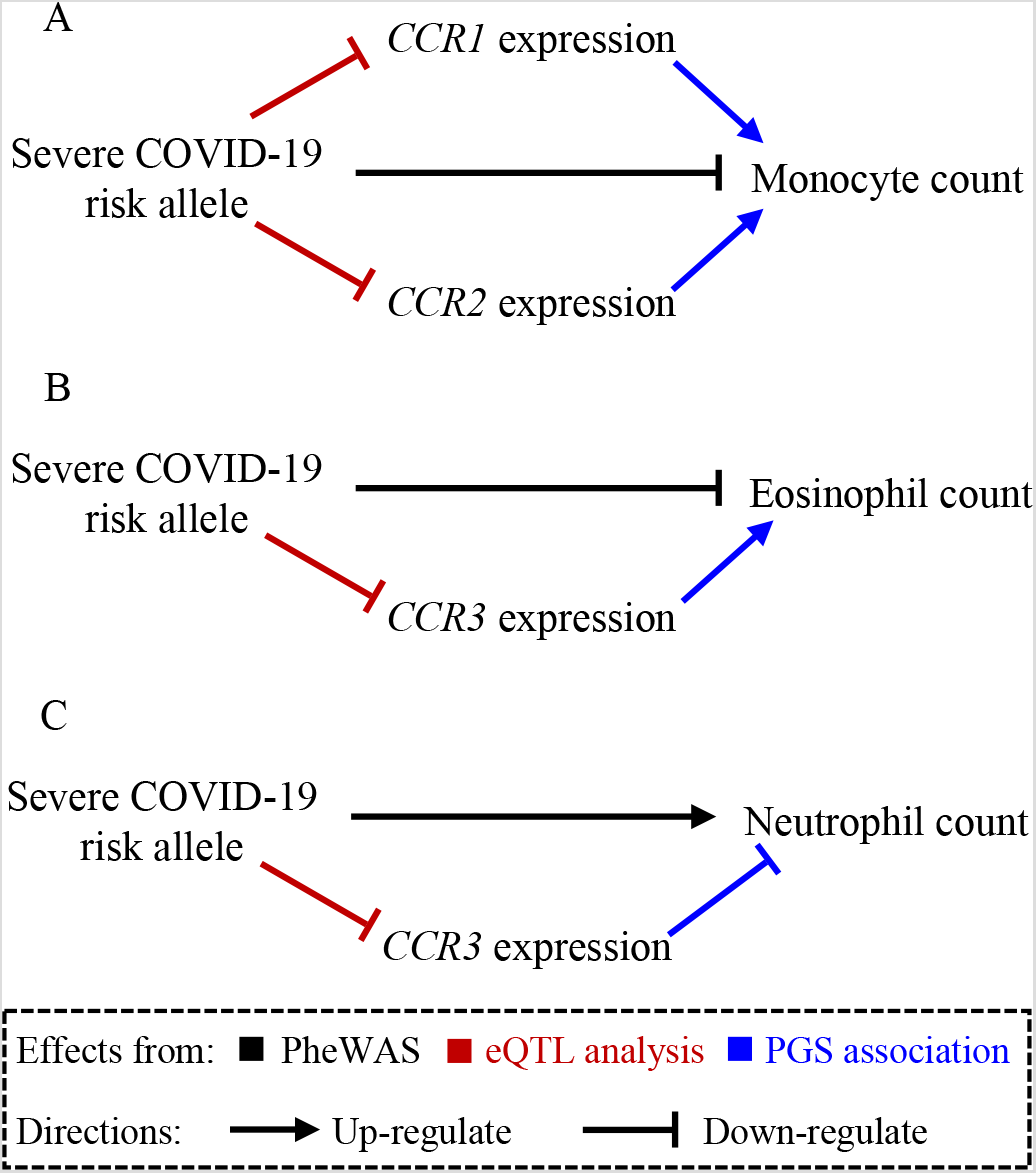
Schematics of possible pathways between the severe COVID-19 risk variant and three blood cell traits. (A) monocyte count, (B) eosinophil count, and (C) neutrophil count. The directions of effects, either up-regulating or down-regulating, were inferred from PheWAS, eQTL analysis in blood samples, and PGS association analysis.

## DISCUSSION

With an unbiased phenome-wide scan approach, our study established two pairs of relationships: 1) associations of the severe COVID-19 risk variant with blood cell traits; and 2) associations between expression levels of candidate target genes and the PGS of blood cell traits. Integrating association signals across multiple analyses prioritizes three blood cell traits, the counts of monocyte, eosinophil and neutrophil, and their candidate target genes, *CCR1, CCR2*, and *CCR3*. Taken together, our results proposed blood cell traits as the probable mechanistic link between the risk variant at 3p21.31 and severe COVID-19.

Hematologic manifestations are common in COVID-19 patients, especially elevated WBC and neutrophil counts but decreased lymphocyte and platelet counts^22,23^. Leukocytosis, neutrophilia, lymphopenia, thrombocytopenia, and neutrophil-to-lymphocyte ratio have been repeatedly associated with worse COVID-19 outcomes and could serve as prognostic biomarkers^1,3,24,25^.Reducing basophil count or percentage was generally observed in patients^26,27^. For monocyte, its total number in circulation does not change dramatically in COVID-19 patients^22,27,28^. However, its composition exhibits a pronounced shift, with a significant expansion of its inflammatory subsets, which are not typically seen in healthy individuals^22,23,28–30^. The pattern of eosinophil is less well-established. Some studies observed diminished and even undetectable eosinophil counts (i.e., eosinopenia) in COVID-19 patients^26,27,31–34^, and it was also shown that eosinophil counts are positively associated with lymphocyte counts in both severe and non-severe cases^34^. However, others did not find a significant difference^35^, and there is also a report of an expanded eosinophil percentage among the total viable leukocyte CD45+ population^22^. These changes in circulating blood cells are closely related to the infiltration and accumulation of lymphocyte, neutrophil, eosinophil, and inflammatory monocyte-macrophage in the lung and other organs, leading to neutrophil extracellular trap and cytokine release syndrome^23,36–38^. Notably, an immuno-monitoring study of COVID-19 patients from acute to recovery phages observed gradual reduction of neutrophil and replenishment of basophil, eosinophil and non-inflammatory monocyte^39^.

Our PheWAS in UK Biobank for the severe COVID-19 risk variant, with replications in another dataset, revealed that the risk allele is associated with decreased monocyte count and percentage, eosinophil count and percentage, but with increased neutrophil percentage. GeneATLAS reported even more significant associations for these relationships, probably due to its different quality control procedures and a larger sample size^14^. It also reports suggestive evidence of negative associations between the risk allele and basophil count and percentage (Supplementary Table 4). These association directions are consistent with the observed blood cell count changes in COVID-19 patients, as discussed above. Of note, our associations were identified in the generally healthy population samples. On the other hand, the vast majority of existing studies measured blood cell counts at hospital admission or during hospitalization, which likely reflect immune responses to SARS-CoV-2 infection. Future studies are warranted to evaluate if before-infection differences in blood cell counts play a role in modulating the risk of developing severe COVID-19.

Our PGS analysis for the potential target genes of the severe COVID-19 risk variant unraveled associations with multiple blood cell counts. It is important to stress that these associations are consistent across analyses with PGS calculated with different GWAS datasets, *p-*value cutoffs, and sample ancestries (Supplementary Table 7). Intersecting and reconciling association signals across PheWAS, eQTL, and PGS analysis yielded multiple possible pathways for the severe COVID-19 risk allele. Strong and consistent evidence was found on the pathways through monocyte and eosinophil. On the other hand, support for the role of neutrophil is weaker. While the association with neutrophil percentage was replicated, the association with neutrophil count was only significant in UK Biobank. The negative associations between *CCR3* expression and PGS of the neutrophil count were only suggestive (*p* = 0.011). In addition to these three blood cells, basophil may serve as another candidate pathway: the risk allele downregulates *CCR3* expression, reduces its stimulatory effect on basophil count, and thus leads to a reduction of basophil. Additional evidence for the potential importance of these candidate genes could be drawn from their cell-type-specific expression patterns (Supplementary Figure 1). *CCR3* has highly specific expression in eosinophil and basophil and only slight expression in neutrophil, *CCR2* has high expression in basophil and medium expression in classical monocyte, while *CCR1* has medium to high expression across all types of granulocytes and monocyte. Notably, this pathway prioritization analysis utilized eQTL association signals in blood samples, but the regulatory effects could be different across tissues^17^. Also, the eQTL analyses were based on generally healthy samples^10,17^. The regulatory effects of the risk variant may be different under the SARS-CoV-2 infection. Further studies are needed to examine its functional effects in patients and to identify the most relevant tissue. Nonetheless, our study prioritized hematologic processes as the downstream pathophysiology of a major genetic locus of severe COVID-19.

The strengths of our study include the unbiased phenome-wide approach at two levels of analysis, the genetic variant (923 phenotypes) and the gene expression (1,263 phenotypes). The large sample sizes in UK Biobank (N = 310,999) and eQTLGen (N = 31,684) increase the statistical power to identify associations. The two phenome-wide analyses converged on multiple blood traits and the association directions are consistent with existing studies in COVID-19 patients. This study also has some weaknesses. First, there are still phenotypes not covered in our analyses, such as monocyte subsets, T cells and B cells. It is possible that the severe COVID-19 risk variant affects other unexplored intermediate traits. Moreover, future fine-mapping studies are needed to identify causal variants at locus 3p21.31 for both severe COVID-19 and blood cell traits. The possibility could not be ruled out at present that they are different variants in strong LD^40^. Our analyses were restricted to population samples and may not reflect patterns in COVID-19 patients. Additionally, the associations between gene expression and genetically predicted blood cell counts should be further confirmed with direct analysis of their measured counts.

In conclusion, our phenome-wide association study for the severe COVID-19 risk variant at locus 3p21.31 and its candidate target genes identified altered blood cell traits, especially counts of monocyte, eosinophil, and neutrophil, as the probable mechanistic links between the genetic locus and severe COVID-19. These blood cell traits, together with their candidate acting genes, *CCR1, CCR2* and *CCR3*, represent compelling and testable hypotheses that call for follow-up studies into their roles in COVID-19 pathogenesis.

## Data Availability

All summary statistics are provided as supplementary materials. Individual-level data could be accessed with approved applications to UK Biobank.

https://www.ukbiobank.ac.uk/

## ACKNOWLEDGES

The authors would like to thank the UK Biobank participants and administrators for data access. We also want to thank all other Ye lab members for stimulating discussions. KY is supported by the University of Georgia Research Foundation. Funding sources had no involvement in the conception, design, analysis, or presentation of this work.

## DISCLOSURE

The authors declare no conflicts of interest.

## SUPPLEMENTARY INFORMATION

**Supplementary Table 1**. PheWAS results for binary disease traits.

**Supplementary Table 2**. PheWAS results for continuous traits, including blood and urine biomarkers, and blood cell traits.

**Supplementary Table 3**. Baseline characteristics of UK Biobank participants.

**Supplementary Table 4**. Significant associations between rs67959919 and blood cell traits in GeneATLAS.

**Supplementary Table 5**. Significant SNP-gene expression associations for rs67959919 (G/A) in GTEx.

**Supplementary Table 6**. Significant SNP-gene expression associations for rs67959919 (G/A) in eQTLGen.

**Supplementary Table 7**. Associations between gene expression and the polygenic scores of blood cell counts. Significant p and FDR values are highlighted.

**Supplementary Figure 1**. The expression of *CCR1, CCR2*, and *CCR3* across blood cells. Figures were retrieved from the Human Protein Atlas (https://www.proteinatlas.org/). The y axis, NX, stands for normalized expression level.

## Notes

### Competing Interest Statement

The authors have declared no competing interest.

### Author Declarations

UK Biobank was approved by the North West Multi-Centre Research Ethics Committee.

